# Fluoxetine pharmacokinetics and tissue distribution suggest a possible role in reducing SARS-CoV-2 titers

**DOI:** 10.1101/2020.12.17.20248442

**Authors:** Andy R. Eugene

## Abstract

**Background:** Recent *in vitro* studies have shown fluoxetine inhibits the severe acute respiratory syndrome coronavirus 2 (SARS-Cov-2) pathogen and one clinical study reported fluoxetine exposure at a median dose of 20mg in patients with the SARS-Cov-2 coronavirus disease 2019 (COVID-19) had a significantly lower risk of intubation and death. The aim of this study is to conduct *in silico* population dosing simulations to quantify the percentage of patients achieving a trough level for the effective concentration resulting in 90% inhibition (EC90) of SARS-Cov-2 as reported in Calu-3 human lung cells.

**Methods:** Population pharmacokinetic parameter estimates for a structural one-compartment model with first-order absorption was used to simulate fluoxetine concentration-time data. A population of 1,000 individuals were simulated at standard fluoxetine doses (20mg/day, 40mg/day, and 60mg/day) to estimate the percentage of the patients achieving a trough level for the EC90 SARS-Cov-2 inhibitory concentration at each day throughout a 10-day treatment period. All analyses were conducted via statistical programming in R.

**Results:** Standard fluoxetine antidepressant doses resulted in a range of 79% to 97% of the patient population achieving a *trough* target plasma concentration of 25.1 ng/ml which translates to lung-tissue distribution coefficient of 60-times higher (EC90 of 4.02 μM). At a dose of 40mg per day, at least 85% of patients will reach the *trough* target EC90 concentration within 3-days. The findings of this pharmacokinetic dosing study corroborate both *in vitro* and *observational* clinical study findings showing fluoxetine inhibits the SARS-Cov-2 pathogen at commonly treated doses in the practice of psychiatry.

## Introduction

The selective serotonin reuptake inhibitor (SSRI) fluoxetine is a racemic mixture of two stereoisomers, (*R)*-fluoxetine and (*S)*-fluoxetine, and maintains regulatory approvals for a wide-array of clinical indications in the practice of psychiatry. Two recent, *in vitro*, studies showed fluoxetine inhibits replication of the Severe Acute Respiratory Coronavirus-2 (SARS-Cov-2) pathogen (Schloer et al., 2020; Zimniak et al., 2020). Specifically, Zimniak et al. reported that following a 3-day incubation period of fluoxetine in Vero cells, inoculated at a multiplicity of infection (MOI) of 0.5, resulted in the median maximal effective concentration (EC50) of 387 ng/ml (1.1 μM) and further found a concentration of 800 ng/ml (2.3 μM) significantly inhibited SARS-Cov-2 replication (Zimniak et al., 2020). Similarly, Schloer *et al*. found that fluoxetine significantly decreases SARS-Cov-2 titers, after a 48-hour incubation period, in both African green monkey kidney epithelial Vero E6 cells (EC50 = 0.69 μM and 90% maximal effective concentration [EC90] = 1.81 μM, MOI=0.01) and human-lung Calu-3 cells (EC50 = 0.82 μM and EC90 = 4.02 μM, MOI=0.1) (Schloer et al., 2020). Taken together, these *in vitro* studies prove in a dose-dependent manner, the SSRI fluoxetine inhibits the SARS-CoV-2 pathogen known to cause the worldwide pandemic, the novel coronavirus disease 2019 (COVID-19).

Considering the well-established clinical symptoms of COVID-19 affecting the lungs, fluoxetine lung concentrations would be an important factor to consider when interpreting any study results. Johnson *et al*. reported human-tissue concentrations, in airline pilots, of fluoxetine in whole-blood ranged from 0.021-1.4 μg/ml and lung concentrations ranged from 1.56 μg/ml to 51.9 μg/ml, leading to a fluoxetine distribution coefficient of 60 (Johnson, Lewis & Angier, 2007). Clinically, the fluoxetine SARS-Cov-2 *in vitro* findings were corroborated by Hoertel *et al*. who showed in a multicenter observational retrospective cohort study of patients who were treated with fluoxetine and diagnosed with COVID-19, experienced a lower risk of intubation and death (hazard ratio=0.32; 95% confidence interval, 0.14-0.73, p=0.007) at a median fluoxetine dose of 20mg (standard deviation [SD]=4.82) (Hoertel et al., 2020). In this context, the aim of this study is to conduct *in silico* population pharmacokinetic dosing simulations to quantify the percentage of patients expected to achieve the *trough* effective concentration resulting in 90% inhibition of SARS-Cov-2.

## Materials & Methods

### Pharmacokinetic Model

Pharmacometric model estimates for differential equation parameters and respective variances for a structural one-compartment pharmacokinetic model with first-order absorption were used to simulate fluoxetine concentration-time data. Model estimates were derived from drug plasma concentrations in 25 females taking a mean dose of 29.4 mg (7.5-80 mg/day) when fluoxetine plasma levels were at steady-state due to being collected for analysis at a minimum median time of fluoxetine treatment of greater than 40-days (Tanoshima et al., 2014). The following parameters were used: volume of distribution (Vd) value of 20.5 liters (variance [ω], 1.24), clearance rate (CL) value of 13.3 liters/hour (ω=0.052), and absorption rate (Ka) of 0.016 (1/hour) (ω=0.231) (Tanoshima et al., 2014).

### Target fluoxetine plasma concentration to achieve EC90 lung concentration

The molecular weight of fluoxetine hydrochloride is 345.8 g/mol and the reported EC50 (0.82 μM) and EC90 (4.02 μM) values from the Schloer *et al*. study is equivalent to: EC50 = 283.6 ng/ml and EC90 = 1390.1 ng/ml, respectively. With the whole-blood to plasma fluoxetine concentration – that is the fraction unbound in plasma – being 0.94 and having whole-blood to lung distribution coefficient of 60, the EC50 and EC90 values would need to be 6% higher than the aforementioned EC50 and EC90 results to then scale the final expected plasma concentration to 1/60 that of the final lung concentration (Sommi, Crismon & Bowden, 1987; Johnson, Lewis & Angier, 2007). For all calculations, the *trough* target plasma concentration is referenced from the Schloer *et al*. study who reported after a 48-hour incubation period in Calu-3 lung cells the 90% maximal effective concentration is 4.02 μM (Schloer et al., 2020) which is significantly higher than the EC90 in Vero E6 cells (1.81 μM) and EC50 results from Zimniak *et al*. and the Schloer *et al*. studies (Schloer et al., 2020; Zimniak et al., 2020).

### Dosing Simulations

To estimate the percentage of patients from a population of one thousand simulated patients who would achieve the *trough* target EC90 concentration, pharmacokinetic dosing of fluoxetine will consist of three dosing trials of fluoxetine: 20mg/day, 40mg/day, and lastly 60mg per day.

### Software and Statistics

All pharmacokinetic dosing simulations are conducted with a population of 1,000 patients using *mrgsolve* and pharmacokinetic parameter estimates using *PKNCA* in R (R Core Team, 2015). Statistical results providing percentage estimates are calculated from trough concentrations of patients achieving the effective concentrations and is referenced from the Schloer *et al*. study reporting the EC90 value in human-lung Calu-3 cells. The whole-blood to plasma ratio of fluoxetine is 0.94 due to being 94% bound to plasma proteins (Sommi, Crismon & Bowden, 1987).

## Results

The fraction of fluoxetine unbound in human plasma is 94%, which requires a 6% increase in plasma levels to achieve whole-blood concentrations to subsequently approximate the blood:lung tissue distribution ratio of 1:60 (Johnson, Lewis & Angier, 2007). Therefore, the new target fluoxetine lung concentration decreasing the SARS-Cov-2 titers by 90% is 1473.5 ng/ml (1390.1 [original, 4.02 μM] + 83.4 ng/ml [6% of original]) and 1/60 of this concentration is the new EC90-plasma concentration of 25.1 ng/ml. The percentage of the 1,000 simulated patients achieving the *trough* EC90-plasma level of 25.1 ng/ml are illustrated in **Figure 1** (20mg/day), **Figure 2** (40mg/day), and **Figure 3** (60mg/day) with a horizontal dashed line throughout the pharmacokinetic dosing profile figures throughout the text and tables.

**Figure 1:**
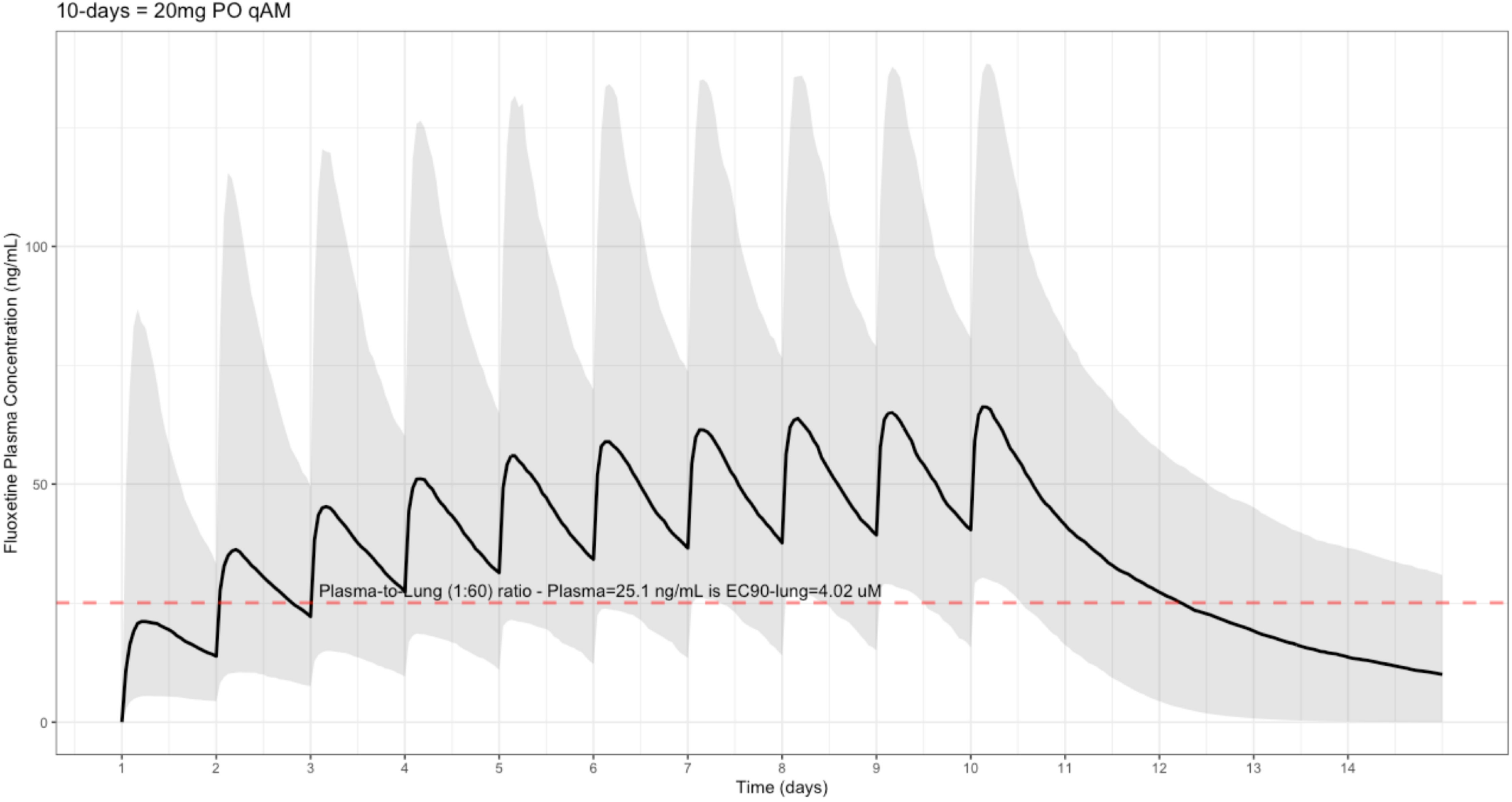
Fluoxetine population (n=1,000) dosing simulation results for an oral dose of *20mg/day* for 10-days. The shaded regions illustrate the 10th (lower) and 90th (upper) percentiles with the solid line within the shaded region representing the median fluoxetine concentration. The dashed horizontal line depicts the effective concentration resulting in 90% inhibition (EC90) of SARS-Cov-2 that will result in 60-times higher level in the lungs.

**Figure 2:**
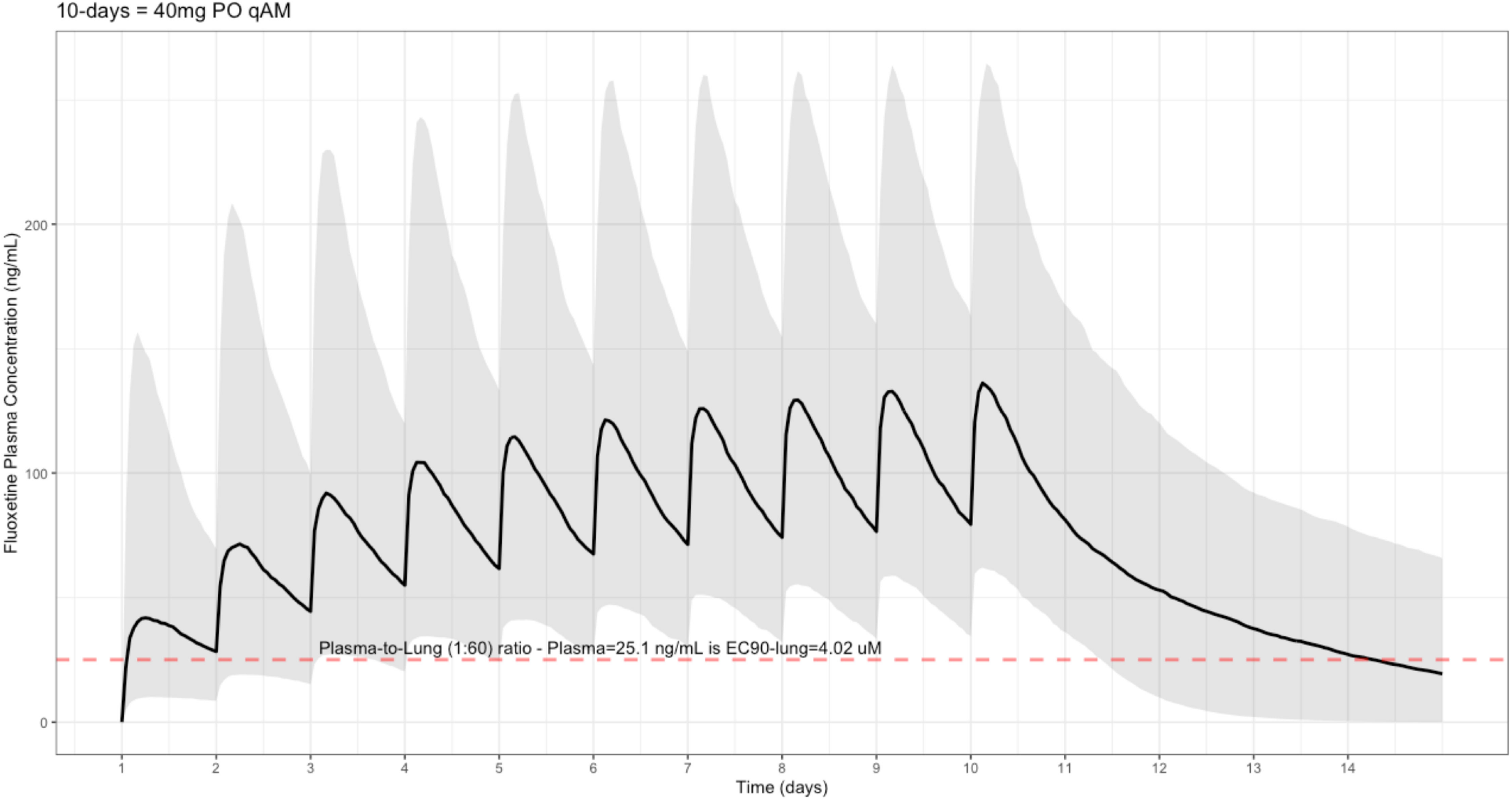
Fluoxetine population (n=1,000) dosing simulation results for an oral dose of *40mg/day* for 10-days. The shaded regions illustrate the 10th (lower) and 90th (upper) percentiles with the solid line within the shaded region representing the median fluoxetine concentration. The dashed horizontal line depicts the effective concentration resulting in 90% inhibition (EC90) of SARS-Cov-2 that will result in 60-times higher level in the lungs.

**Figure 3:**
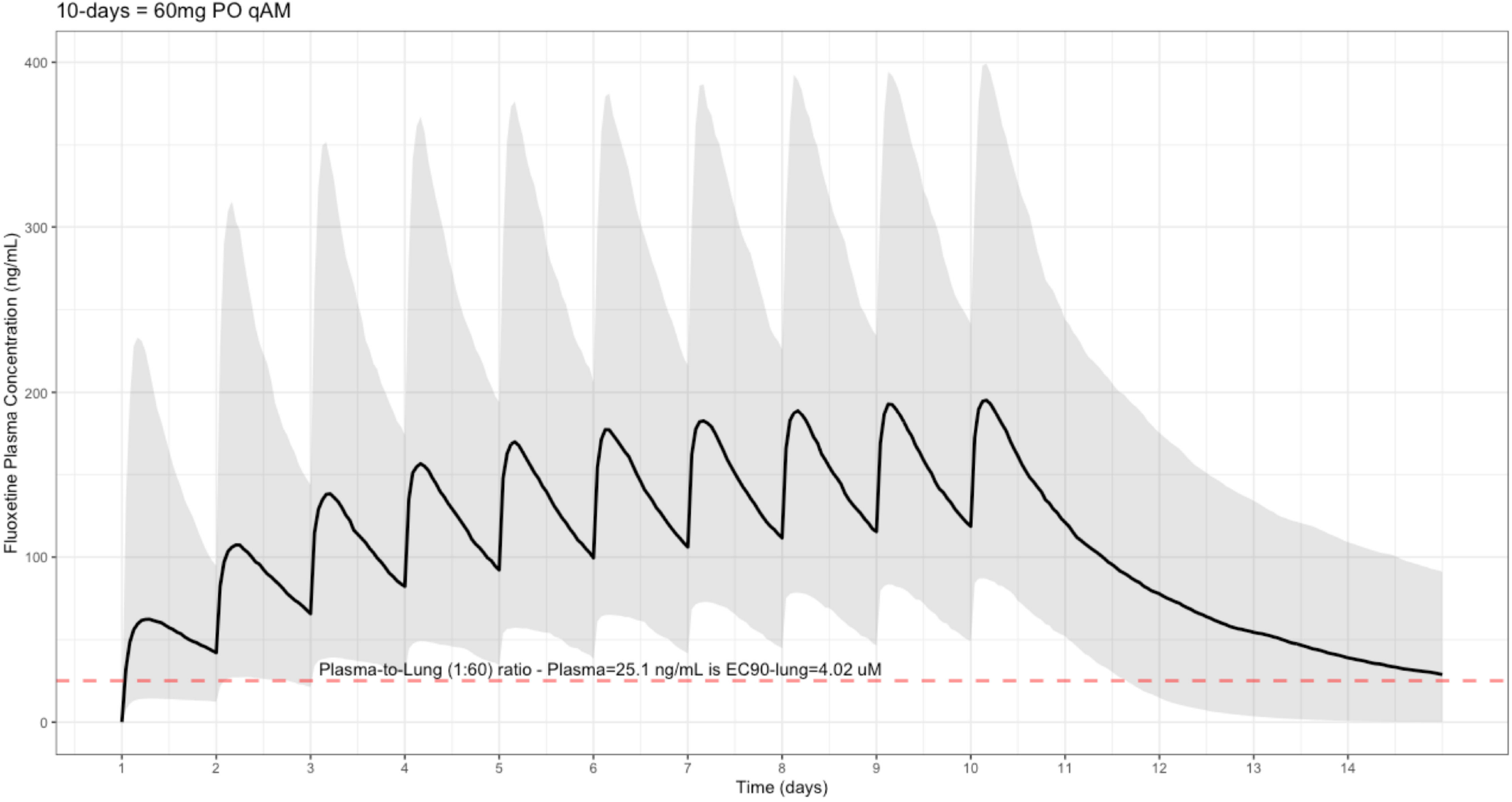
Fluoxetine population (n=1,000) dosing simulation results for an oral dose of *60mg/day* for 10-days. The shaded regions illustrate the 10th (lower) and 90th (upper) percentiles with the solid line within the shaded region representing the median fluoxetine concentration. The dashed horizontal line depicts the effective concentration resulting in 90% inhibition (EC90) of SARS-Cov-2 that will result in 60-times higher level in the lungs.

**Figure 1** shows the concentration-time data for a fluoxetine dose of 20mg per day results in the maximum plasma concentration (Cmax), geometric mean (geometric coefficient of variation, CV%), is 65.8 ng/mL (CV=70.2%), median time at maximum concentration (Tmax) of 220-hours (range, 49-220), area-under-the-concentration-time curve (AUC_0→Last_), geometric mean (geometric coefficient of variation, CV%), from baseline to 10-days is 10,200 ng•hour/ml, and a half-life (t ½) – expressed as arithmetic mean (standard deviation, SD) – of 84.7 hours (SD=181). These aforementioned pharmacokinetic results translate to 20% of the population reaching the target concentration at the end of day-1 and 79% of the population achieving the target *trough* EC90 concentration by end of day-10. **Figure 2** shows at a dose of 40mg per day, the Cmax is 132 ng/mL (CV=68%), Tmax of 220-hours (range, 49-220), AUC_0→Last_ is 20,500 ng•hour/ml, and population t ½ is 81.4 (SD=113), which is interpreted as 55% of the population achieving the EC90 *trough* target at day-1 and 95% by day-10. Moreover, **Figure 3** shows in a patient population treated with fluoxetine at 60mg daily, results in a Cmax of 191 ng/mL (CV=71%), 220-hours (range, 49-232), AUC_0→Last_ 29,700 ng•hour/ml, and t ½ of 85.4 (SD=209) allowing 72% of the population reaching the target *trough* concentration threshold on day-1 and 97% by day-10 of fluoxetine treatment. **Table 1** provides an overview of the pharmacokinetics and pharmacodynamics with blood levels (ng/ml and μM) in plasma as well as calculated organ concentrations (whole-blood, lung, brain, heart, liver, spleen, and kidney) as well as, the percent of the population achieving *trough* EC90 target during a treatment period of 10-days.

**Table 1:** Fluoxetine pharmacokinetics and pharmacodynamics showing blood levels (ng/ml and μM) and percent of population achieving plasma trough concentration of 25.1 ng/ml leading to a lung-EC90 target of 4.02 μM during a treatment period of 10-days.

| Fluoxetine concentrations at standard antidepressant doses |  |  |  |
| --- | --- | --- | --- |
|  | 20mg/day | 40mg/day | 60mg/day |
| | Cmax ng/ml [$\mu\text{M}$] | Cmax ng/ml [$\mu\text{M}$] | Cmax ng/ml [$\mu\text{M}$] |
| Plasma | 65.5 [0.19] (CV=70%) | 132 [0.38] (CV=68%) | 191 [0.55] (CV=71%) |
| Whole-Blood | 69.4 [0.20] | 139.9 [0.40] | 202.5 [0.59] |
| Lung | 4165.8 [12.1] | 8395.2 [24.3] | 12147.6 [35.1] |
| Brain | 1041.5 [3.0] | 2098.8 [6.1] | 3036.9 [8.8] |
| Heart | 694.3 [2.0] | 1399.2 [4.1] | 2024.6 [5.9] |
| Liver | 2638.3 [7.6] | 5317.0 [15.4] | 7693.5 [22.3] |
| Spleen | 1388.6 [4.0] | 2798.4 [8.1] | 4049.2 [11.7] |
| Kidney | 624.9 [1.8] | 1259.3 [3.6] | 1822.1 [5.3] |

| Percent of population (n=1,000) achieving *trough* SARS-Cov-2 EC90 target |  |  |  |
| --- | --- | --- | --- |
| Time Point | 20mg/day | 40mg/day | 60mg/day |
| Day 1 | 19.6% | 55.2% | 71.9% |
| Day 2 | 42% | 77.2% | 87.1% |
| Day 3 | 56.2% | 85.6% | 91.4% |
| Day 4 | 62.5% | 89.3% | 93.9% |
| Day 5 | 68% | 91.5% | 95.1% |
| Day 6 | 71.5% | 92.9% | 95.6% |
| Day 7 | 74.3% | 93.5% | 96.2% |
| Day 8 | 76.3% | 94% | 96.5% |
| Day 9 | 77.8% | 94.2% | 96.7% |
| Day 10 | 78.9% | 94.6% | 96.8% |

EC90: effective concentration inhibiting 90% of SARS-Cov-2 from Calu-3 lung cells reported as 4.02 $\mu\text{M}$ (Schloer et al., 2020). Maximum plasma concentration (Cmax) are expressed as geometric mean (geometric coefficient of variation, CV%). Fluoxetine tissue distribution coefficients (60 for lung, 20 for spleen, 15 for brain, 10 for heart, and 9 for kidneys) were scaled from whole-blood, then plasma, and then to organs as reported (Johnson, Lewis & Angier, 2007).

## Discussion

According to the United States Food and Drug Administration (FDA) Adverse Events Reporting System (FAERS) during the window period of 1982 to June 30, 2020, fluoxetine was reported to have a total of 79,929 cases, 62,948 serious cases, and 10,043 end of life cases (United States Food and Drug Administration, 2018). Females represented 58% of the adverse drug reactions (ADRs), males represent 27% of the ADRs, and 15% of the ADRs did not specify a gender. The most common adverse drug event reported for fluoxetine is *Drug Interaction* and amounts to 3,798 cases (4.75% of total). Given this information, drug interactions associated with fluoxetine are due to inhibition of the cytochrome P450 (CYP) system, specifically CYP2C19 and CYP2D6 which may have interactions such as in patients taking tamoxifen in oncology – by inhibiting conversion to the active endoxifen metabolite via CYP2D6 – or in cases of clopidogrel – inhibiting the conversion to the active 2-oxo-clopidogrel metabolite – in cardiology. (Spina, Trifirò & Caraci, 2012; Eugene, 2019; United States Food and Drug Administration, 2020).

Extrapolating from *in vitro* to *in vivo* concentrations are dependent on intracellular versus extracellular concentrations, as well as methodology of quantifying either whole-blood versus plasma concentrations in human pharmacokinetic studies. The EC50 and EC90 target concentrations represent the extracellular fluoxetine concentrations in the SARS-Cov-2 cell culture media. As COVID-19 is known to affect the brain during active infection and in post-COVID-19 states, adequate brain concentrations would be clinical importance in patients who may experience depression. Bolo *et al*. reported fluoxetine brain concentrations, at steady-state, using fluorine magnetic spectroscopy showed fluoxetine concentrations were 10-times higher in the brain than in human plasma (Bolo et al., 2000). Specifically, Bolo *et al*. found in study participants taking oral doses (10mg, n=1; 20 mg, n=1; 40 mg, n=2) with a treatment period ranging from 3-months to 12-months, had fluoxetine human brain concentrations of 13 μM (SD=7) versus 1.73 μM (SD=1.00) in human plasma fluoxetine (Bolo et al., 2000). In comparison, Johnson *et al*. found the coefficients for tissue distribution of fluoxetine relative to whole-blood was: 60 for lung, 20 for spleen, 15 for brain, 10 for heart, and 9 for kidneys (Johnson, Lewis & Angier, 2007).

As patients recover the from the acute COVID-19 symptoms, long-term sequelae are being documented as a term coined as: post-COVID-19 syndrome. In one of the post-SARS-Cov-2 infection studies in young patients, 92% were found to have ongoing cardiorespiratory symptoms with organ dysfunction representing comprised of: 32% impairment of the heart, 33% impairment of lungs, and 12% impairment of the kidneys (Dennis et al., 2020). In another post-COVID-19 syndrome study, 96% of the patients were female and experienced statistically significant exercise intolerance, dyspnea, and chest pain when compared to those not diagnosed with COVID-19 (Walsh-Messinger et al., 2020). Moreover, Walsh-Messinger *et al*. study found patients with post-COVID-19 syndrome had higher ratings of depression subscale markers of altered sleep and thinking, but depression severity was not significantly different with patients not diagnosed with COVID-19 (Walsh-Messinger et al., 2020).

Direct clinical translation of this current pharmacokinetic study is corroborates with a retrospective multicenter observational study, by Hoertel *et al*., who found a median fluoxetine dose of 20mg resulted in a significantly lower risk of intubation and death in a population composed of 63% women and 37% men (Hoertel et al., 2020). Moreover, Hoertel *et al*. found 17% of fluoxetine-treated patients had events of intubation and death appears consistent with the current study due to as simulations are extended to 4-weeks at 20mg 82.9% of patients would achieve the EC90 target and 17% would not achieve the EC90 concentration. Comparing the Hoertel *et al*. and Zimniak *et al*. publications, Hoertel *et al*. found that in addition to fluoxetine, venlafaxine (median dose of 75mg) and escitalopram (median dose of 10mg) were also associated with a lower risk of intubation and death, however, Zimniak *et al*. showed neither SSRIs escitalopram nor paroxetine inhibited SARS-Cov-2 *in vitro* (Hoertel et al., 2020; Zimniak et al., 2020). Of note, as shown in **Table 1**, a 40mg or 60mg daily fluoxetine dose results in 90% inhibition of the SARS-Cov-2 infection due to surpassing the EC90 value of 4.02 μM as found in Calu-3 cells and the EC90 value of 1.81 μM in Vero E6 cells (Schloer et al., 2020).

Antiviral properties of fluoxetine are well reported in the literature. A study by Carpinteiro *et al*. reported fluoxetine inhibits acid sphingomyelinase preventing infection of both cultured cells and human nasal epithelial cells in SARS-Cov-2 as well as in vesicular stomatitis virus pseudoviral particles presenting the SARS-CoV-2 spike protein (Carpinteiro et al., 2020). A study by Zuo *et al*. showed fluoxetine resulted in potent inhibition of the coxsackievirus by reducing both synthesis of viral RNA and protein with an EC50 of 2.3 μM and peak antiviral properties at 6.25 μM (Zuo et al., 2012). A study by Bauer *et al*. showed, in a broad-spectrum manner, fluoxetine inhibited enterovirus – the picornaviridae family – replication with the (*S)*-fluoxetine enantiomer exhibiting a 5-fold lower EC50 than the racemic mixture of (*R)-* and (*S)*-fluoxetine (Bauer et al., 2019). Further, Bauer *et al*. found the following EC50 values: coxsackievirus B3 (racemate-EC50=2.02 μM, (*S*)-fluoxetine-EC50=0.42 μM), enterovirus EV-D68 (racemate-EC50=1.85 μM, (*S*)-fluoxetine-EC50=0.67 μM), and (*S*)-fluoxetine values alone for rhinovirus HRV-A2 (EC50=7.95 μM) and HRV-B14 (EC50=6.34 μM) (Bauer et al., 2019). Notably, Zimniak *et al*. found that individual stereoisomers, (*R)*-fluoxetine and (*S)*-fluoxetine, inhibited the SARS-Cov-2 viral load; however, in contrast, fluoxetine could not inhibit gene expression of the herpes simplex-1 virus, human herpes virus-8, rabies virus, nor the respiratory syncytial virus (Zimniak et al., 2020). Lastly, of these aforementioned antiviral median effective concentrations, standard fluoxetine doses achieve the concentrations in the lungs, brain, and heart, except for the rhinovirus EC50 for the heart tissue but which will be reached in the brain and lungs, as shown in **Table 1**.

A limitation of this study is associated with the previously validated fluoxetine pharmacometric model being in women and did not include men (Tanoshima et al., 2014). However, as shown from the aforementioned FDA Adverse Events Reporting System data, women represented 58% of all ADR cases overall from the reporting period of 1982 to 2020. Overall, from a drug-safety perspective, prior to administering fluoxetine, a careful review of all patient medications and clinical status by clinical pharmacologist-physicians would be recommended to avoid drug interactions due to fluoxetine’s ability to strongly inhibit CYP2C19 and CYP2D6 (Hefner, 2018). Compounds that are sensitive and moderate CYP2C19 (e.g. omeprazole diazepam, lansoprazole, rabeprazole, voriconazole) and CYP2D6 substrates (e.g. dextromethorphan, eliglustat, nebivolol, tolterodine, encainide, metoprolol, propranolol, tramadol) will have an increase total area-under-the-concentration-time curve of ≥ 5-fold drug exposure when treated with fluoxetine (United States Food and Drug Administration, 2020). Lastly, patients who have a pharmacogenomic profile of being a CYP2D6 Poor Metabolizer or Intermediate Metabolizers should be closely monitored for potential fluoxetine side-effects, but may also have a higher rate of achieving the target trough EC90 concentration at a 20mg daily fluoxetine dose relative to CYP2D6 Normal (Extensive) Metabolizers.

## Conclusions

Investigating fluoxetine pharmacokinetics, this study confirmed that previously published median effective concentrations and specifically the EC90 fluoxetine value inhibiting SARS-Cov-2 in Calu-3 human lung cells is achievable using standard fluoxetine doses (20mg/day, 40mg/day, and 60mg/day) and also corroborates findings from a retrospective clinical study showing fluoxetine exposure associated with reduced risk of intubation and death. Due to the high fluoxetine brain (15-times higher) and lung (60-times higher) tissue distributions, relative to whole-blood and plasma concentrations (6% lower than whole blood), treatment with fluoxetine may serve as pragmatic therapeutic option in patients with lingering post-COVID-19 syndrome neurocognitive effects or in primary management of COVID-19. Overall, assuming patients are not treated with medications that result in drug interactions with fluoxetine – that is are sensitive or moderate CYP2D6 metabolic substrates – a dose of 40mg per day of fluoxetine will likely be most effective with inhibiting SARS-Cov-2 titers. That 40mg daily fluoxetine dose will lead to 55% of the population achieving the trough EC90 target at day-1, 93% by day-7, and 95% of patients achieving the trough target EC90 concentration within 10-days of fluoxetine.

## Data Availability

The study is based on simulated pharmacokinetic data.

## Acknowledgements

None.

